# Characteristic Fetal Brain MRI Abnormalities in Pyruvate Dehydrogenase Complex Deficiency

**DOI:** 10.1101/2024.04.08.24303574

**Authors:** Olivier Fortin, Kelsey Christoffel, Abdullah Shoaib, Charu Venkatesan, Kate Cilli, Jason W. Schroeder, Cesar Alves, Rebecca D. Ganetzky, Jamie L. Fraser

## Abstract

Pyruvate dehydrogenase complex deficiency (PDCD) is a disorder of mitochondrial metabolism that is caused by pathogenic variants in multiple genes, including *PDHA1*. Typical neonatal brain imaging findings in PDCD have been described, with a focus on malformative features and chronic encephaloclastic changes. However, fetal brain MRI imaging in confirmed PDCD has not been comprehensively described. We sought to demonstrate the prenatal neurological and systemic manifestations of PDCD determined by comprehensive fetal imaging and genomic sequencing.

All fetuses with a diagnosis of genetic PDCD who had undergone fetal MRI were included in the study. Medical records, imaging data, and genetic testing results were reviewed and reported descriptively.

Ten patients with diagnosis of PDCD were included. Most patients had corpus callosum dysgenesis, abnormal gyration pattern, reduced brain volumes, and periventricular cystic lesions. One patient had associated intraventricular hemorrhages. One patient had a midbrain malformation with aqueductal stenosis and severe hydrocephalus. Fetuses imaged in the second trimester were found to have enlargement of the ganglionic eminences with cystic cavitations, while those imaged in the third trimester had germinolytic cysts.

Fetuses with PDCD have similar brain MRI findings to neonates described in the literature, although some of these findings may be subtle early in pregnancy. Additional features, such as cystic cavitations of the ganglionic eminences, are noted in the second trimester in fetuses with PDCD, and these may represent a novel early diagnostic marker for PDCD. Using fetal MRI to identify these radiological hallmarks to inform prenatal diagnosis of PDCD may guide genetic counseling, pregnancy decision-making, and neonatal care planning.

## Introduction

Pyruvate dehydrogenase complex (PDC) deficiency (PDCD) is an inborn error of metabolism caused by pathogenic variants in various genes related to the PDC, a multiple enzyme complex that plays a critical role in mitochondrial energy production.^1–3^ Pyruvate, a product of glycolysis, is normally converted by the PDC into acetyl-CoA, which then enters the citric acid cycle to produce ATP. When PDC cannot convert pyruvate to acetyl co-A, the excess pyruvate is converted into lactate, causing lactic acidosis and energy failure that are often initial signs of PDCD. For this reason, the ketogenic diet is often used to bypass carbohydrate metabolism and the subsequent energy failure (Figure 1).^1,3–5^

**Figure 1:**
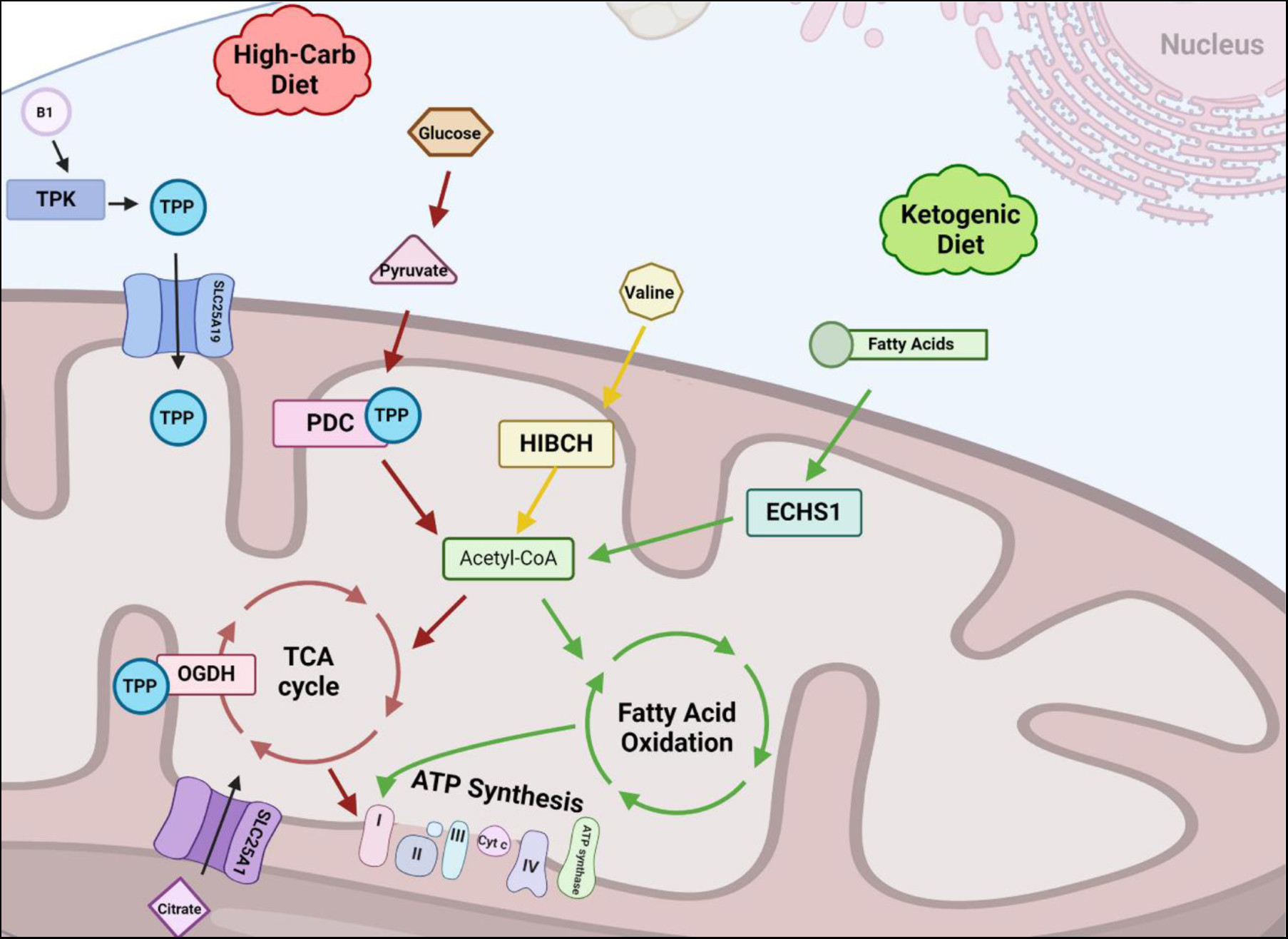
Schematic representation of the PDC and related enzymatic pathways. Thiamine pyrophosphate is the requisite cofactor for the PDC as well as branched chain amino acid catabolism and tricarboxylic acid cycle function. Ketogenic diet bypasses the PDCD complex while enabling ATP synthesis through fatty acid oxidation. ATP: adenosine triphosphate; B1: vitamin B1; ECHS1: short-chain enoyl-CoA hydratase; HIBCH: beta-hydroxyisobutyryl-CoA hydrolase; PDC: pyruvate dehydrogenase complex; OGDH: oxoglutarate dehydrogenase; TCA: tricarboxylic acid; TPK: thiamine pyrophosphokinase; TPP: thiamine pyrophosphate. Created with BioRender.com.

The most common pathogenic variants associated with PDCD occur in *PDHA1* which encodes the alpha subunit of the pyruvate dehydrogenase enzyme (a component of the E1 subenzyme of the PDC).^1,3,6^ Inheritance for this gene is X-linked, but both males and females have been described with *PDHA1* pathogenic variants. Neonates with the severe neurologic disease are usually female.^7^ Rare variants (all with autosomal recessive inheritance) have also been identified in genes coding other components of the PDC, including *PDHB*, *DLAT*, *DLD*, *PDHX*, and the regulatory enzyme *PDP1*.^1,3^ PDCD also occurs secondary to conditions that impair PDC function, such as dietary thiamine deficiency and enzyme dysfunction upstream in the biochemical pathway that lead to deficiencies in cofactors necessary for PDC function.^3^ Examples of such genetic disorders include Thiamine pyrophosphokinase deficiency (related to biallelic pathogenic variants in *TPK1*), Mitochondrial short-chain enoyl-CoA hydratase 1 deficiency (related to biallelic pathogenic variants in *ECHS1*), and 3-hydroxyisobutyryl-CoA hydrolase deficiency (related to biallelic pathogenic variants in *HIBCH*).

Multiple phenotypes of PDCD occur with onset in neonates, infants, children, and even adults.^1,8,9^ The metabolic phenotype of encephalopathy, lactic acidosis, hypotonia, and seizures commonly present in neonates with PDCD with or without central nervous system malformations.^3,10,11^ Craniofacial abnormalities are common, including microcephaly, frontal bossing, low-set ears, upturned nose, long flat philtrum, and thin upper lips.^1,3^ Although there is no curative treatment for PDCD, ketogenic diet and thiamine supplementation have been described to improve neurological outcomes in mild to moderate cases.^3,10^ Severe cases may show little benefit from treatment, and there is significant risk of death in the first few months of life. Typical brain MRI findings of neuronopathic PDCD have been documented in infants and children. These include chronic encephaloclastic changes – periventricular white matter cavitations, diffuse parenchymal atrophy, and ventriculomegaly – as well as corpus callosum dysgenesis and brainstem malformations.^1,3,6,10,12^ A lactate peak on MR spectroscopy (MRS) has also been described in neonates with PDCD.^5,13,14^ Children with PDCD may also present later in life with Leigh-like changes on brain MRI, including T2-weighted changes and diffusion restriction involving the basal ganglia – with particular predilection for the globus pallidus – as well as the thalamus and brainstem.^10,12,15^ Congenital brain malformations may not be present in childhood-onset PDCD.

Imaging findings of PDCD in the prenatal period are less well described, with few case reports and series of brain abnormalities identified prenatally, either on fetal ultrasound or MRI. Imaging findings of neuronopathic PDCD in the prenatal period may include corpus callosum dysgenesis, cortical gyration abnormalities, ventriculomegaly, reduced white matter volumes, as well as reduced brainstem and cerebellar volumes.^6,13,16–20^ One series reported fetal MRI findings for 3 fetuses who were diagnosed with PDCD, one with pathogenic variants in *PDHA1* and two (siblings) with pathogenic variants in *PDHB*.^6^ Fetal MRS, although currently limited in its clinical application,^21–23^ was reported to be normal in a fetus with suspected PDCD (based on family history and clinical course but not confirmed with genetic testing), with no lactate peak.^16^

Given the paucity of data on fetal imaging and diagnosis in this rare disorder, we sought to report cases of PDCD identified through fetal diagnostic clinics in pediatric hospital centers in the United States.

## Materials and Methods

Retrospective medical records review was performed to identify fetuses and neonates with a genetic diagnosis of PDCD who underwent fetal imaging by MRI between January 1, 2010, and October 17, 2023 in the fetal diagnostic clinics at the authors’ institutions. All authors obtained either IRB approval or IRB exemption from their respective centers. Waiver of consent was granted based on the retrospective nature of the study.

Available medical records, imaging studies, and genetic testing of the fetus and mother were reviewed. Postnatal medical records were reviewed, if available. Pregnancy history, maternal medical history, family history, imaging features, genetic testing results, and clinical outcomes were analyzed and reported in a descriptive fashion. To further de-identify these cases, at the direction of the journal’s editorial office, the gestational ages (GA) of all cases are indicated within two-week GA ranges throughout.

Fetal MRI imaging for the cohort was overread by fetal and pediatric neuroradiologist J.W.S. for descriptive and technical consistency. Fetal imaging at Children’s National Hospital was performed on a 1.5 Tesla scanner (GE HealthCare, Chicago, Illinois; Siemens Healthineers, Erlangen, Germany); unenhanced fast T1-weighted, T2-weighted, and diffusion-weighted images were generated in various planes based on the local fetal imaging protocol. Imaging at Cincinnati Children’s Hospital Medical Center was performed on a 1.5 Tesla scanner (GE HealthCare, Chicago, Illinois; Philips Healthcare, Best, Netherlands). Imaging at Children’s Hospital of Philadelphia was performed on a 1.5 Tesla scanner (Siemens Healthineers, Erlangen, Germany). Imaging performed at University of Texas Southwestern was performed on a 1.5 Tesla scanner (Siemens Healthineers, Erlangen, Germany).

Neonatal imaging at Children’s National Hospital was performed on a 1.5 Tesla scanner (GE HealthCare, Chicago, Illinois). Unenhanced fast T1-weighted, T2-weighted, diffusion-weighted, and susceptibility-weighted images were generated in orthogonal planes based on the local neonatal imaging protocol; when done, MR spectroscopy was obtained using single voxel long and short TE through the left basal ganglia. Imaging at Cincinnati Children’s Hospital Medical Center was performed on a 1.5 Tesla or 3 Tesla scanner (GE HealthCare, Chicago, Illinois; Philips Healthcare, Best, Netherlands). Imaging at Children’s Hospital of Philadelphia was performed on a 1.5 Tesla scanner (Siemens Healthineers, Erlangen, Germany). Imaging at University of Texas Southwestern was performed on a 1.5 Tesla scanner (Siemens Healthineers, Erlangen, Germany).

Genetic testing strategies varied at the discretion of the treating providers at the time of the clinical evaluation and included single-gene testing, multi-gene panels, and exome sequencing (ES). Hospital laboratories or commercial laboratories (MNG Laboratories, Atlanta, Georgia; GeneDx, Gaithersburg, Maryland; Claritas Genomics, Cambridge, Massachusetts) were used based on patient insurance coverage, institutional policies, and availability of testing in the respective centers. Genetic variant pathogenicity was reported by clinical genetics laboratories using recognized ACMG criteria; they were individually evaluated for confirmation and or diagnostic validity by three of the authors (O.F., R.D.G., J.L.F.).

## Results

### Demographics

Ten cases of diagnosed PDCD who underwent fetal MRI in fetal diagnostic clinics at the authors’ institutions were included. All cases were genetically confirmed to be female. Six cases were evaluated between 20w and 29w gestational age (GA), whereas 4 fetuses were evaluated between 30w and 35w GA (Table 1).

**Table 1:** Genetic variants, fetal MRI findings, and outcomes in reported patients with PDCD.

| **<u>Case</u>** | **<u>Gene</u>** | **<u>Variant and ACMG Classification</u>** | **<u>Referral Indication</u>** | **<u>Fetal MRI Findings</u>** | **<u>Pregnancy Outcome</u>** |
| --- | --- | --- | --- | --- | --- |
| **1** | *PDHA1* | c.252_253delGA<br>p.K85Nfs*10<br>Pathogenic | Bilateral VM | 24-26w GA: Microcephaly, severe VM, delayed sulcation, dysplastic CC, small cerebellum, bilateral GE cystic cavitations | Pregnancy continued |
| **2** | *PDHA1* | c.951_961del<br>p.M318Gfs*18<br>Likely pathogenic | Borderline VM, periventricular calcifications | 24-26w GA: VM, delayed sulcation, hypogenesis of the CC, small volumes, unilateral GE cystic cavitations | Pregnancy continued |
| **3** | *PDHA1* | c.788G>C<br>p.R263P<br>Pathogenic | Absent CC, VM | 20-22w GA: Complete ACC, decreased volumes, bilateral GE cystic cavitations | Pregnancy continued |
| **4** | *PDHA1* | c.904C>T<br>p.R302C<br>Pathogenic | Mild VM, declining HC | 28-30w GA: Near-complete ACC, mild-moderate VM, decreased volumes, delayed sulcation, bilateral GE cystic cavitations | Termination of pregnancy |
| **5** | *PDHA1* | c.831G>A<br>p.K277=<br>Likely pathogenic | Absent CSP and CC | 20-22w GA: Near-complete ACC, bilateral GE cystic cavitations | Termination of pregnancy |
| **6** | *PDHA1* | c.934_940del<br>p.S312Vfs*12<br>Pathogenic | Severe VM | 30-32w GA: severe VM, dysplastic CC, cerebellar and vermis hypoplasia, abnormal sulcation, bilateral germinolytic cysts | Pregnancy continued |
| **7** | *PDHA1* | c.385del<br>p.L129Ffs*51<br>Pathogenic | Bilateral VM | 28-30w GA: cerebellar and vermis hypoplasia, brainstem hypoplasia, aqueductal stenosis, severe VM | Pregnancy continued |
| **8** | *PDHA1* | c.968_976del<br>p.R323_V325del<br>Likely pathogenic | Severe VM, abnormal cortex | 34-36w GA: severe VM, bilateral IVH, bilateral germinolytic cysts, subependymal heterotopias, PMG | Termination of pregnancy |
| **9** | *PDHA1* | c.1154_1164del11<br>p.K385fs*43<br>Pathogenic | Bilateral VM, PFM | 30-32w GA: dysplastic CC, cerebellar hypoplasia, mild VM, subependymal periventricular heterotopias, periventricular and intraventricular cysts | Pregnancy continued |
| **10** | *TPK1* | c.246C>A; p.Y82*<br>c.365T>C; p.I122T<br>Likely pathogenic | Absent CSP, microcephaly | 22-24w GA: complete ACC, microcephaly, reduced brain volumes, periventricular cysts, and bilateral GE cystic cavitations | Termination of pregnancy |
| ACC: agenesis of the corpus callosum; CBLH: cerebellar hypoplasia; CC: corpus callosum; CCD: corpus callosum dysgenesis; CSP: cavum septum pellucidum; GA: gestational age; GE: ganglionic eminence; HC: head circumference; IVH: intraventricular hemorrhage; MCD: malformation of cortical development; PFM: posterior fossa malformation; PMG: polymicrogyria; PV: periventricular; VM: ventriculomegaly |  |  |  |  |  |

### Fetal Imaging

Fetal MRI findings are detailed in Table 1 and Figure 2. Eight fetuses had abnormalities of corpus callosum (either ACC or dysplastic CC), all cases had parenchymal volume loss, 7 cases had varying degrees of ventriculomegaly, 5 cases had cerebellar hypoplasia, and 6 cases had some form of cortical malformation (either delayed sulcation or frank polymicrogyria). Nine cases had cystic changes: those imaged in the second trimester or early third trimester (n=6) had cystic cavitations in one or both ganglionic eminences (GE), whereas those imaged later in the pregnancy (n=3) had periventricular or germinolytic cysts. One fetus imaged between 28 weeks and 30 weeks GA (Case 7) had a midbrain malformation with aqueductal stenosis that resulted in severe and progressive ventriculomegaly; cystic changes were not seen in this fetus. Figure 3 and 4 show examples of fetal and neonatal brain imaging.

**Figure 2:**
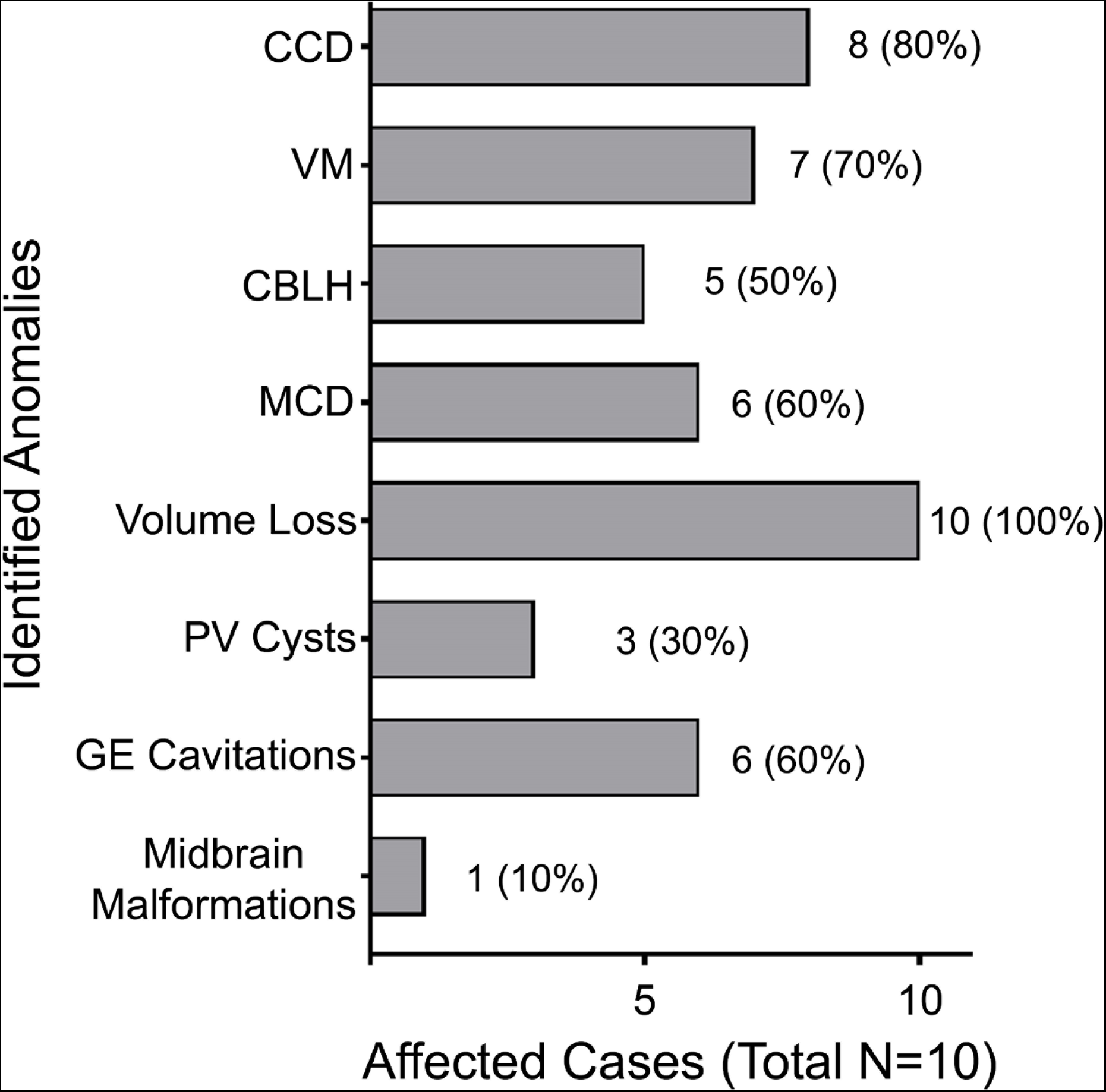
Fetal MRI Findings in Fetuses with PDCD. Most fetal cases had multiple brain anomalies or malformations at the time of fetal MRI (N=10). CBLH: cerebellar hypoplasia; CCD: corpus callosum dysgenesis; GE: ganglionic eminence; MCD: malformation of cortical development; PV: periventricular; VM: ventriculomegaly

**Figure 3:**
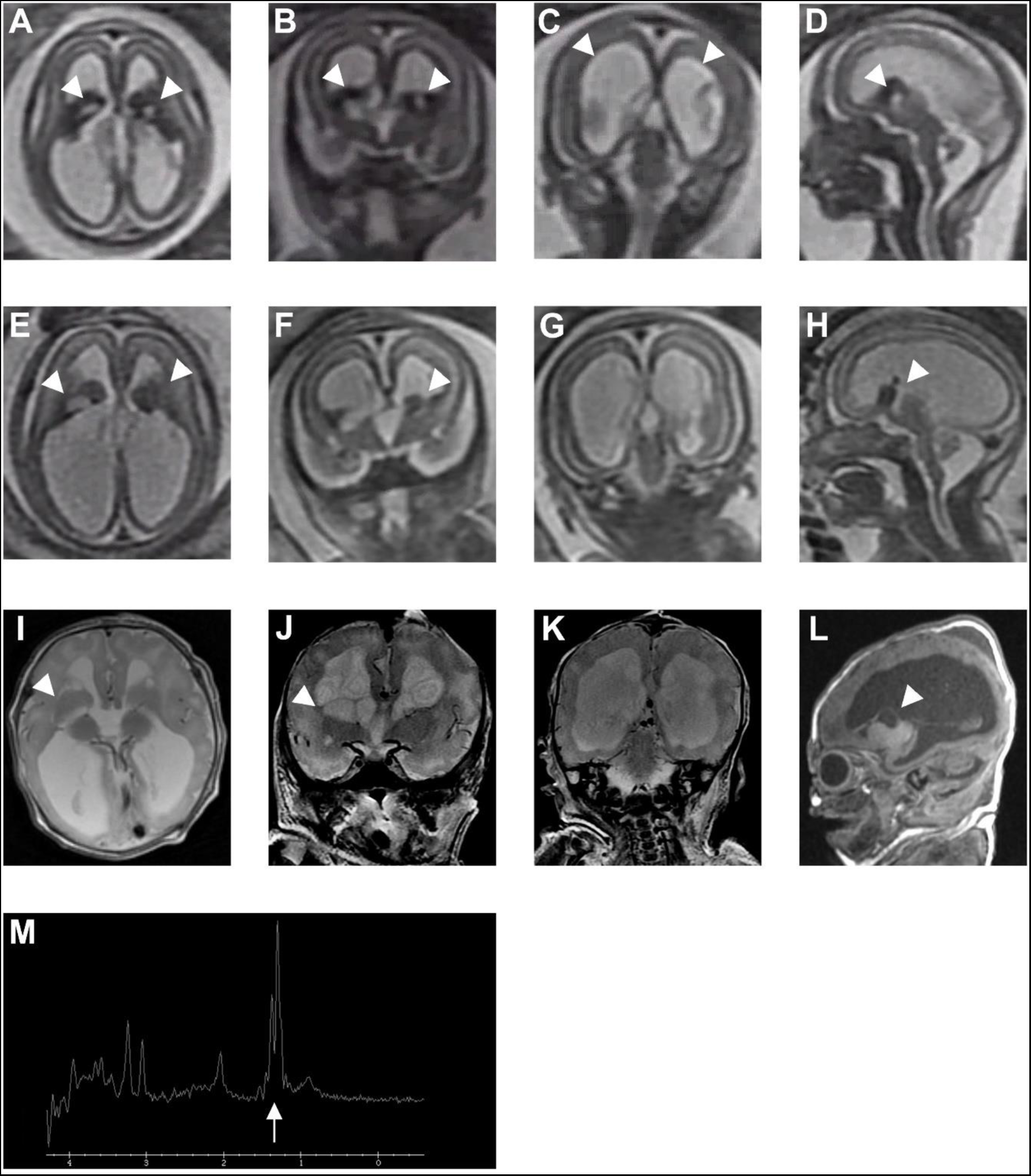
Fetal and postnatal brain MRI/MRS findings in PDCD (Case 1) Imaging findings from Case 1 include the first prenatal imaging (20-22w GA; **A**-**D**), the second prenatal imaging (24-26w GA; **E**-**H),** and neonatal imaging with magnetic resonance spectroscopy (**I**-**L)**. **A**-**D**: T2-weighted fetal MRI in the axial (**A**), coronal (**B**, **C**), and sagittal (**D**) planes, showing ganglionic eminence cystic cavitations (while arrowheads; **A**, **B**, **D**), ventriculomegaly (white arrowhead; **C**), and corpus callosum dysgenesis. **E**-**H**: T2-weighted fetal MRI at in the axial (**E**), coronal (**F**-**G**), and sagittal (**H**) planes, showing progression of ganglionic eminence changes (arrowheads; **E**, **F**, **H**) and ventriculomegaly (arrowheads; **G**), as well as corpus callosum dysgenesis. **I**-**L**: T2 weighted (**I**, **J**, **K**) and T1-weighted (**L**) neonatal brain MRI in the axial (**I**), coronal (**J**, **K**), and sagittal (**L**) planes, showing basal ganglia abnormalities with surrounding cysts (arrowheads; **I**, **J**, **L**), progression of ventriculomegaly (arrowheads; **K**), volume loss, and corpus callosum dysgenesis. **M**: Neonatal brain MRS showing elevated lactate peak (arrow).

**Figure 4:**
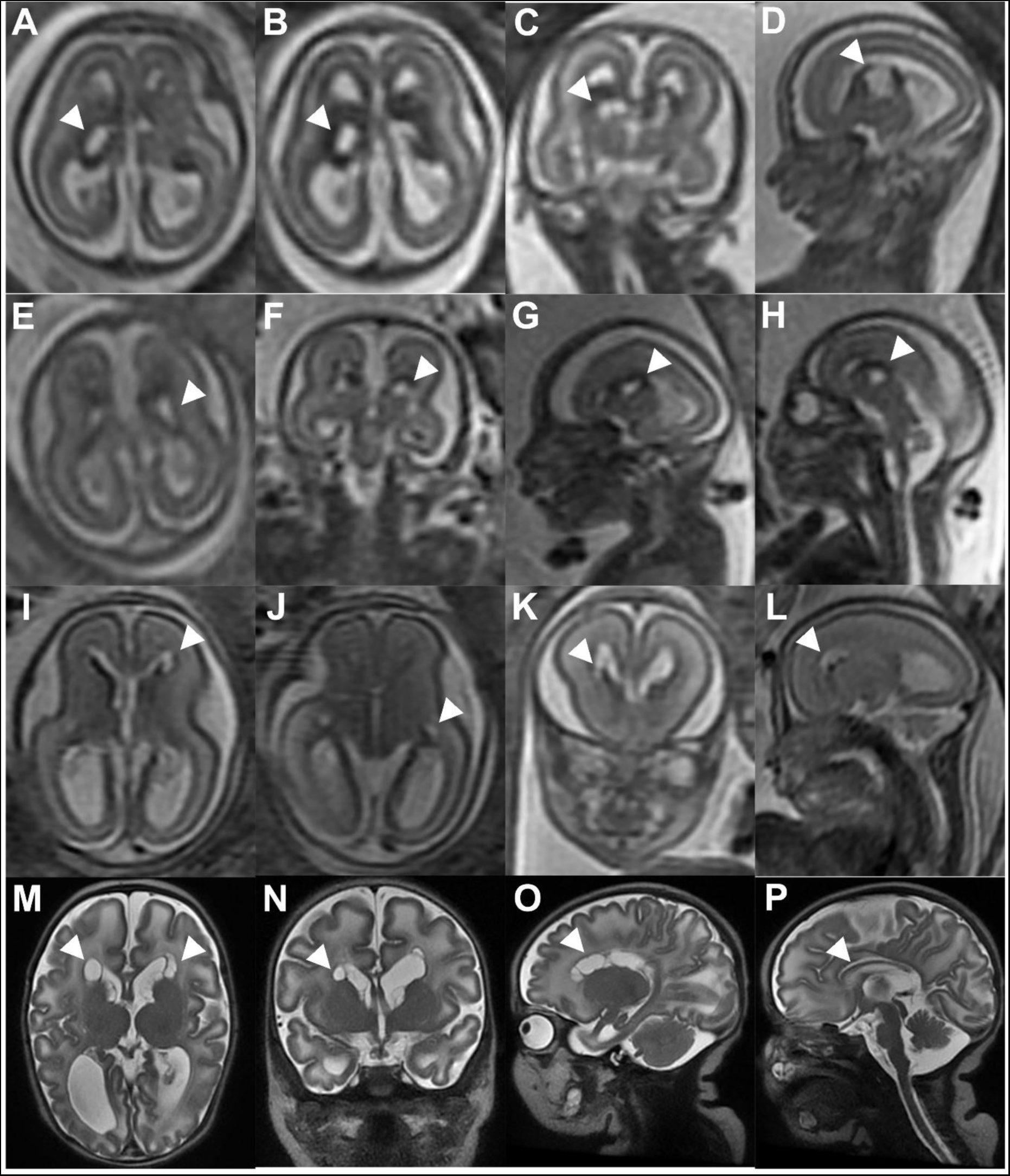
Fetal and postnatal brain MRI findings in select cases of PDCD. **A-D**: T2-weighted fetal MRI (20-22w GA) in the axial (**A**, **B**), coronal (**C**), and sagittal (**D**) planes in a fetus with *PDHA1*-related PDCD (Case 5), showing ganglionic eminence cystic cavitations (arrowheads; **A**, **B**, **C**, **D**) and corpus callosum dysgenesis. **E**-**H**: T2-weighted fetal MRI (22-24w GA) in the axial (**E**), coronal (**F**), and sagittal (**G**-**H**) planes in a patient with *TPK1*-related disorder (Case 10), showing ganglionic eminence cystic cavitations (arrowheads; **E**, **F**, **G**, **H**) and corpus callosum dysgenesis. **I**-**L**: T2 weighted fetal MRI at (24-26w GA) in the axial (**I**, **J**), coronal (**K**), and sagittal (**L**) planes in a patient with *PDHA1*-related PDCD (Case 2), showing periventricular cysts (arrowheads; **I**, **J**, **K**, **L**). **M**-**P**: T2-weighted postnatal brain MRI in the axial (**M**), coronal (**N**), and sagittal (**O**-**P**) planes in a patient with *PDHA1*-related PDCD (Case 2), showing progression of the periventricular cysts (arrowheads; **M**, **N**, **O**) and hypoplastic corpus callosum (arrowhead; **P**).

### Genetic Diagnoses

Three patients underwent amniocentesis for genetic testing by ES; all results were obtained postnatally or after termination. Two patients underwent genetic testing from products of conception. Five patients underwent genetic testing in the postnatal period.

Nine cases were eventually found to have heterozygous pathogenic or likely pathogenic variants in *PDHA1*, consistent with a primary PDCD deficiency (Table 1). One case was found to have biallelic (compound heterozygous) likely pathogenic variants in *TPK1*.

### Pregnancy Outcomes

Termination of pregnancy occurred in 4 cases based on the poor neurological prognosis. The remaining cases (n=6) were delivered at term, except for 1 who was delivered late preterm (between 34 weeks and 36 weeks GA). Neonatal management and postnatal outcomes are detailed in Table 2. Two infants expired in the neonatal intensive care unit (Cases 1 and 7). Ketogenic diet was started during the neonatal period in Cases 3 and 6. Case 9 was started on ketogenic diet in infancy for infantile spasms. For Case 2, ketogenic diet was recommended but not started by the family. Five patients underwent follow-up brain MRI in the neonatal period (Table 2). Three neonates also underwent brain MRS.

**Table 2:** Postnatal Management and Outcomes.

| <u>Case</u> | <u>Gestational Age</u> | <u>Neonatal Management</u> | <u>Neonatal MRI Findings</u> | <u>Postnatal Outcome</u> |
| --- | --- | --- | --- | --- |
| **1** | 37-39w | NICU admission for severe neonatal encephalopathy and lactic acidosis | Dysplastic CC, CBLH, brainstem hypoplasia, severe VM, PV cysts, dysgyria<br>MRS: elevated lactate peak | Profound metabolic acidosis unresponsive to all intensive care interventions attempted. Natural death occurred on DOL 3. |
| **2** | 34-37w | Normal delivery and neonatal course. | Dysplastic CC, mild VM, PV cysts<br>No MRS | Seen in outpatient clinic. Motor delays in infancy. Ketogenic diet recommended but not pursued. |
| **3** | 37-39w | NICU admission for neonatal encephalopathy and lactic acidosis. Started on ketogenic diet and thiamine. | Hypoplastic CC, vermis hypoplasia, moderate VM, PV cysts<br>MRS: normal | Discharged home on thiamine and ketogenic diet. |
| **6** | 37-39w | NICU admission for neonatal encephalopathy, lactic acidosis, hyperammonemia, and seizures. Treated with levetiracetam and ketogenic diet. | Complete ACC, severe VM, microcephaly, abnormal white matter signal.<br>MRS: large lactate doublet | Discharge home on levetiracetam and ketogenic diet. |
| **7** | 37-39w | NICU admission for management of severe hydrocephalus and respiratory failure | Hypoplastic Z-shaped brainstem, CBLH, aqueductal stenosis, severe VM, complete ACC, thin cortical mantle<br>MRS: large lactate doublet | Natural death occurred in neonatal period. |
| **9** | 37-39w | NICU admission for mild neonatal encephalopathy and apneic episodes | Complete ACC, CBLH, severe VM, PV cysts, white matter volume loss<br>No MRS | Discharged home. Later presented with infantile spasms and started on ketogenic diet. |
| ACC: agenesis of the corpus callosum; CBLH: cerebellar hypoplasia; CC: corpus callosum; DOL: day of life; GA: gestational age; GE: ganglionic eminence; HC: head circumference; NICU: neonatal intensive care unit; PV: periventricular; VM: ventriculomegaly |  |  |  |  |

## Discussion

We present ten cases of PDCD evaluated in comprehensive prenatal diagnosis programs for abnormal fetal brain findings and later found to have pathogenic variants in genes either within the PDC or in upstream or adjacent enzymatic pathways: nine had de novo heterozygous likely pathogenic or pathogenic variants *PDHA1* and one had biallelic likely pathogenic variants in *TPK1*. Although the brain imaging findings of neonatally-identified PDCD are likely related to prenatal onset of the disease, fetal neuroimaging descriptions are less well described with a lack of correlation with the postnatal features. In this study, we demonstrate the progression of brain malformations from the prenatal to the postnatal period and report on abnormalities previously underappreciated in PDCD.

Although *PDHA1*-related disorder is a recognized cause of congenital brain malformations,^1,10,12^ significant brain malformations have never been reported with *TPK1* disease-causing variants. *TPK1* encodes for the enzyme thiamine pyrophosphokinase, which synthesizes thiamine pyrophosphate (TPP), an essential cofactor for PDC function. *TPK1*-related disorder usually presents in childhood with variable severity of ataxia, dystonia, spasticity, developmental delay, encephalopathy, and hypotonia; these children may also have episodes of neurological decompensation and developmental regression after febrile illness with brain MRI changes reminiscent of Leigh disease.^24–28^ Supraphysiological supplementation of thiamine with or without ketogenic diet has been reported to salvage the metabolic encephalopathic phenotype and improve developmental outcomes.^24–28^ The patient with the youngest reported onset of symptoms in the literature was 1 month of age when she started to have seizures, although the neurological deterioration only presented later in early childhood.^28^ Fetal onset of neurological concerns in *TPK1*-related disorder has not been described in the literature, although a neonatal case reported by two co-authors is forthcoming. The case reported in this study would be the first fetal case of *TPK1*-related disorder. Thus, TPK deficiency may be an underrecognized cause of congenital brain malformations. TPK is highly intolerant to variation in humans, as biallelic loss of function variants have not been observed in humans. Severe TPK deficiency may lead to embryonic lethality, while moderate deficiency may be sufficient to cause the characteristic features of PDCD on fetal MRI. All genes encoding enzymes in the PDC pathway should be evaluated in cases of fetal brain malformations reported herein.

Some of the features on fetal MRI in our patients overlap with the neonatal findings described in the literature,^1,10,12^ including corpus callosum dysgenesis, abnormal gyration pattern, and reduced volumes of the supratentorial and infratentorial compartments of the brain with periventricular cystic cavitations. A brainstem malformation was noted in one patient, resulting in aqueductal stenosis and severe obstructive hydrocephalus, making the diagnosis of a metabolic disorder such as PDCD appear unlikely until an extensive genetic evaluation was performed after birth. One patient had bilateral intraventricular hemorrhages, which also may steer clinicians away from investigating a genetic disorder such as PDCD.

When comparing with the available literature on fetal brain MRI findings in PDCD,^6,13,14,16–20^ we noted that the majority of our cases (90%) had cystic changes, whereas this is reported to be present in about 50% of cases in a recent review.^20^ Cystic changes were not seen in one fetus (Case 7) but these may have been effaced by the severe hydrocephalus. Non-hemorrhagic caudothalamic groove cysts, or germinolytic cysts, were seen in fetuses that were imaged in the third trimester. The other fetuses that were imaged in the second trimester or early third trimester had more prominent cystic cavitations in one or both ganglionic eminences (GE), usually associated with enlargement of these structures. Upon review of the available fetal MRIs of two patients with *PDHA1*- and *PDHB*-related PDCD, reported by Pirot et al.^6^, their imaging between 26 weeks and 28 weeks GA also appears to show cystic cavitations within the GEs, although these were described as “paraventricular pseudocysts” at the time.

GEs are a normal fetal structure within the telencephalon that naturally evolve during central nervous system (CNS) development, later giving rise to the basal ganglia.^29–31^ Based on recent reports of volumetric measurements from quantitative fetal MRI, they appear to increase in size throughout the first and second trimester, then involute in the late second trimester and third trimester.^32^ The GE is divided between medial, lateral, and caudal segments (MGE, LGE, and CGE, respectively), and each segment is thought to contain specific progenitor cells that give rise to various interneurons essential to brain function.^30,33^ For example, cells in the MGE produce GABAergic neurons that project to the striatum and globus pallidus as well as interneurons that migrate tangentially to the cerebral cortex, whereas cells in the CGE produce GABAergic interneurons that project to the limbic system, and cells in the LGE produce interneurons destined for to the olfactory bulb, amongst others.^30^ Complex gene and protein functions control the proliferation, differentiation, and migration of cells arising from GE subsegments.^30^ The GE, as well as pathology involving these structures, may be visualized by fetal ultrasound and MRI.^34–37^ GE cystic cavitations are a rare finding on fetal brain MRI and were first reported in 2013. These findings were initially linked to microlissencephalies related to *LIS1* or *TUBA1A* pathogenic variants.^37,38^ A larger study of 22 fetuses with GE abnormalities, including cystic cavitations, using ES found that this specific malformation was associated with pathogenic variants in *TUBA1A* in 7 patients and *PDHA1* in one patient.^39^ The fetus with PDCD described by Goergen et al.^39^ was imaged between 22 weeks and 24 weeks GA and also had ACC. GE cystic cavitations have not been seen with *TPK1*-related disorder in the literature. Combined with our work, this suggests that cystic cavitations of the GE could be an early diagnostic marker of PDCD on second trimester fetal MRI, especially when associated with corpus callosum dysgenesis; this finding appears to “convert” to the more typical caudothalamic groove or germinolytic cyst in the third trimester. The exact pathophysiological link between PDCD and GE cystic cavitations is unclear. Germinal matrix hemorrhage in premature infants has been shown to alter cell proliferation in the GE via different pathways including inflammation and necrosis^29^; an analogous process may be at play in PDCD, whereby mitochondrial dysfunction and energy failure in this highly proliferative and metabolically demanding structure leads to cell death and subsequent structural changes. Evidence of hemorrhage may be secondary to rapid cell death and involution in this region and not a primary cause of tissue injury. The timing and locations of severe metabolic injury in the fetuses described herein point to the significant energy demand of these developing structures, which is not met in PDCD. The evolution of fetal mitochondrial brain injury in PDCD correlates with observed and expected injury of the most metabolically demanding brain structures postnatally. Thus, mitochondrial dysfunction from PDCD that manifests in the prenatal period disrupts cortical developmental patterning, decreases cerebral volume, and causes tissue destruction of the GE, findings that can be seen on fetal MRI in the second trimester.

Given that the fetal radiological features of *PDHA1*- and *TPK1*-related disorders may mimic destructive and malformation disorders of the CNS, we suggest that all genes causing PDCD be included in the differential diagnosis of many fetal brain disorders, especially with corpus callosum dysgenesis, cortical migration disorders, and periventricular cystic changes in combination. Even in cases with apparently isolated abnormalities of the corpus callosum, one should consider an underlying genetic disorder such as PDCD, given that some of the other changes may not be detected on fetal MRI. Up to 25% of cases with fetal diagnosis of isolated corpus callosum dysgenesis may eventually have a postnatal diagnosis of complex corpus callosum anomaly (i.e. associated with other brain malformations).^40,41^ The yield of expanded genetic testing in isolated and complex agenesis of the corpus callosum is not well known, given that most larger studies have used chromosome microarray (CMA) only; yield of copy number and chromosome-based testing has been reported to be between 5% and 7%.^42–44^ One study reported that 16% of patients with isolated ACC and 26% of patients with complex ACC obtained a genetic diagnosis in the postnatal period, although the testing regimen is not described in detail or standardized.^40^ Given the myriad monogenetic disorders associated with both isolated and complex ACC,^45^ including PDCD, CMA alone is insufficient in most cases. Large gene panels that include *PDHA1* or ES would be necessary to identify these etiologies. The use of ES in prenatal testing for fetal diagnosis has shown use in various neurological and non-neurological fetal phenotypes,^46^ although the yield of systematic sequencing-based testing in fetal diagnosis of ACC is unknown. Given that significant and potentially actionable diagnoses, such as PDCD, that can present with ACC only few other fetal MRI findings – especially early on in fetal brain development, it is essential to offer ES in addition to CMA, although this may depend on local regulatory, insurance, and financial contexts.

While postnatal MRS has been used to support a diagnosis of PDCD and other mitochondrial disorders, fetal MRS is not routinely used. Literature supporting the utility and validity of MRS in the fetal population has recently advanced this concept as a viable modality in prenatal imaging.^21–23^ In cases where mitochondrial disease is suspected based on fetal imaging phenotypes, MRS may provide additional confirmatory evidence of disease and clarify prognosis for families. Advanced fetal imaging, including MRI with volumetric studies and MRS, may be a useful adjunct for prognosis discussions and counselling while genetic testing is pending.

We thus present ten cases of primary or secondary PDCD that were evaluated prenatally with fetal MRI. All cases showed typical MRI findings of PDCD, including corpus callosum dysgenesis, abnormal gyration pattern, and periventricular cystic lesions, although these were at times subtle on fetal imaging. Ganglionic eminence enlargement with cystic cavitation may represent an early diagnostic marker for severe PDCD on second trimester fetal MRI, although disorders such as tubulinopathies must also be considered in the differential diagnosis and genetic testing approach. Fetuses with PDCD may also present with brain malformations that are not typically consistent with this metabolic disorder, such as aqueductal stenosis and hydrocephalus. Genetic testing must include genes related to PDCD in all cases of corpus callosum dysgenesis, cortical malformation, or cystic changes of the brain. We recommend clinical genome sequencing or exome sequencing in combination with chromosomal microarray as the optimal diagnostic approach in these cases to detect both copy number and single nucleotide variants over gene panels or microarray alone. Timely identification of characteristic imaging features of PDCD is necessary to expedite genetic testing and earlier diagnosis, which in turn may impact pregnancy planning and postnatal outcomes.

## Acknowledgements

We would like to thank our patients and their families for their essential contributions to this work. We would also like to thank all staff and providers that have provided care and expertise to the patients and their families at our respective institutions.

## Data availability

De-identified data will be made available upon reasonable request to the corresponding author.

## Funding

This project relied on institutional salary support for the authors. Dr. Christoffel receives research support for unrelated endeavors from T32HD098066. Dr. Fraser receives research support for unrelated endeavors from U01NS106845-01A1, U54NS115052-01, and U24NS131172-01in addition to philanthropic support from grateful families and generous donors.

## Competing interests

The authors report no competing interests

## Notes

### Competing Interest Statement

The authors have declared no competing interest.

### Funding Statement

This study did not receive any funding.

### Author Declarations

The IRB of Children’s National Hospital waived ethical approval for this work. Waiver of consent was granted based on the retrospective nature of the study. The IRB of Children’s Hospital of Philadelphia waived ethical approval for this work. Waiver of consent was granted based on the retrospective nature of the study. The IRB of Cincinnati Children’s Hospital waived ethical approval for this work. Waiver of consent was granted based on the retrospective nature of the study.

